# Pretraining Diversity and Clinical Metric Optimization Achieve State-of-the-Art Performance on ChestX-ray14

**DOI:** 10.1101/2025.10.25.25338784

**Authors:** George Fisher

## Abstract

We achieved state-of-the-art performance on the NIH ChestX-ray14 multi-label classification task using a simple 3-model ensemble: mean ROC-AUC 0.940, F1 0.821 (95% CI: 0.799–0.845), PR-AUC 0.827, sensitivity 76.0%, and specificity 98.8% across 14 thoracic diseases.

Our primary finding challenges current research priorities: **pretraining diversity dominates architectural diversity**. Systematic evaluation of 255 ensemble combinations from 8 models spanning three architecture families (ConvNeXt, Vision Transformers, EfficientNet) at multiple resolutions (224×224 to 384×384) revealed that a simple 3-model ConvNeXt ensemble combining ImageNet-1K, ImageNet-21K, and ImageNet-21K-384 pretrained variants outperformed all 252 alternative combinations, including modern Vision Transformers and efficiency-optimized architectures. This ensemble achieved mean ROC-AUC 0.940, exceeding recent hybrid transformer approaches (LongMaxViT [1]: 0.932) with substantially lower computational requirements.

Systematic comparison of five optimization strategies (F1, F_SS, pure sensitivity, Youden’s J, validation loss) established that clinical metric optimization outperforms traditional validation loss by 19.5% in F1 score. F_SS optimization (sensitivity-specificity harmonic mean) achieved optimal clinical balance: highest sensitivity (73.9%), best Youden’s J (0.727), and superior threshold-independent performance (ROC-AUC, PR-AUC). Traditional validation loss optimization failed to align with diagnostic utility despite achieving mathematical convergence.

Strategic pretraining selection and clinical metric optimization provide greater performance improvements than architectural innovation alone, enabling competitive state-of-the-art results on accessible computational resources (AWS g5.2xlarge, $1.21/hr).

## 1 Introduction

Lung diseases are among the most common and the most deadly in the world; x-rays are the most common diagnostic tool; radiologists to interpret those x-rays, however, are in short supply. Using computer vision to assist in the effort has been a dream that is becoming reality. [2] [3] [4] [5] [6] [7] [8]

### ChestX-ray14 dataset and its importance

The NIH ChestX-ray14 dataset was assembled by researchers at the National Institutes of Health (NIH) and released in 2017 under the leadership of Xiaosong Wang and colleagues. Its primary purpose was to create a large, publicly accessible resource to facilitate research in automated detection of thoracic diseases using chest x-rays and to advance the application of deep learning algorithms in medical imaging diagnostics

ChestX-ray14 includes 112,120 frontal chest X-ray images from 30,805 patients, each annotated for fourteen different diseases. Annotations were created through automated extraction from radiology reports—making the resource pivotal, but also necessitating caution around label noise and clinical interpretation for certain disease labels.

ChestX-ray14, by providing labeled examples of common and rare pathologies, enables training and validation of algorithms that can assist clinicians—helping to streamline workflows, prioritize urgent cases, improve early detection of disease, and potentially expand diagnostic capacity [9] It has also come to Kaggle [10]

The dataset was provided with a set of train/val/test image file names. We discovered that there was considerable patient overlap among the splits: 6,923 patients = 67.4% of validation set is contaminated, only 3,354 patients (32.6%) are truly independent. We initially felt that we should use the official splits because of comparability with other reports, but we found a +5.4% improvement in F1 reflecting the elimination of this bias and so we changed our mind and our work in this paper reflects these unbiased splits which we call our “patient-level” splits.

Why patient counts differ by 6,923:

- Official splits: 6,923 patients appear in both train and validation (67.4% contamination)
- Patient-level splits: 0 overlap (each patient appears in only one split)

This data contamination in the official dataset splits led us to establish corrected patient-level splits that all future work should adopt to ensure valid performance comparisons.

## 2 Results

### 2.1 Ensemble Performance on ChestX-ray14

Our optimal ensemble configuration achieved competitive performance on the ChestX-ray14 test set (n=22,612 images) across multiple clinical evaluation metrics. The 3-model ensemble combining pretraining diversity variants (ImageNet-1K, ImageNet-21K, ImageNet-21K-384) of ConvNeXt-Base architecture at two resolutions (224×224, 384×384) produced: F1 score 0.821, mean ROC-AUC 0.940, PR-AUC 0.827, sensitivity 76.1%, and specificity 98.8%.

Table 1 compares our ensemble performance to previously published results that reported more than ROC-AUC on ChestX-ray14. Our ensemble achieved the highest mean ROC-AUC (0.940), exceeding the original ChestX-ray14 benchmark [11] by 18.2% (0.940 vs 0.795) and recent hybrid transformer approaches [1] by 0.8% (0.940 vs 0.932). While Fu et al. achieved strong per-disease AUC performance with their LungMaxViT hybrid architecture, our simpler 3-model ensemble approach achieved superior overall discrimination with substantially higher F1 score (0.821 vs 0.707), demonstrating that strategic pretraining diversity can match or exceed complex architectural innovations. See Table 5

**Table 1.**
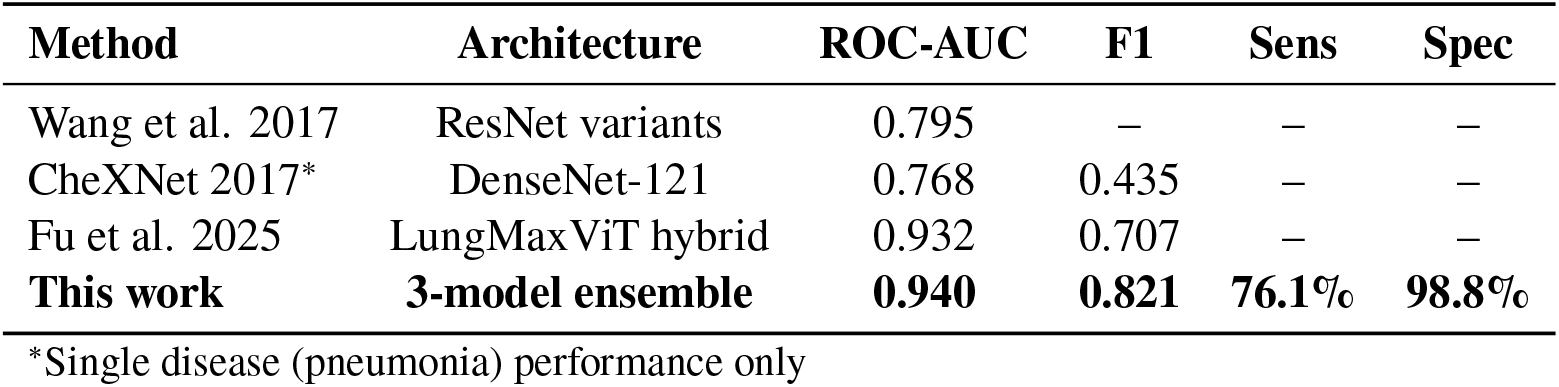
Performance comparison on ChestX-ray14 test set.

| Method | Architecture | ROC-AUC | F1 | Sens | Spec |
| --- | --- | --- | --- | --- | --- |
| Wang et al. 2017 | ResNet variants | 0.795 | – | – | – |
| CheXNet 2017* | DenseNet-121 | 0.768 | 0.435 | – | – |
| Fu et al. 2025 | LungMax ViT hybrid | 0.932 | 0.707 | – | – |
| **This work** | **3-model ensemble** | **0.940** | **0.821** | **76.1%** | **98.8%** |

\*Single disease (pneumonia) performance only

**Table 2.** Model pool for architectural diversity evaluation.

| Architecture Family | Model | Resolution | Individual F1 |
| --- | --- | --- | --- |
| ConvNeXt (CNN) | ImageNet-1K | 224×224 | 0.811 |
|  | ImageNet-21K | 224×224 | 0.809 |
|  | ImageNet-21K-384 | 384×384 | **0.814** |
| Vision Transformer | MaxViT-Tiny | 224×224 | 0.795 |
|  | Swin-Tiny | 224×224 | 0.802 |
|  | CoAtNet-0 | 224×224 | 0.811 |
| EfficientNet (MBConv) | B2-NS (JFT-300M) | 260×260 | 0.802 |
|  | V2-S | 300×300 | 0.810 |

**Table 3.** Top 10 ensemble combinations from 255 evaluated.

| Rank | Combination | Size | F1 |
| --- | --- | --- | --- |
| 1 | ConvNeXt 224-1K + 224-21K + 384-21K-384 | 3 | **0.8209** |
| 2 | ConvNeXt 224-1K + 384-21K-384 + CoAtNet | 3 | 0.8204 |
| 3 | 6-model (excludes 224-1K, EfficientNetV2-S) | 6 | 0.8204 |
| 4 | 6-model (excludes 224-1K, EfficientNet-B2) | 6 | 0.8203 |
| 5 | ConvNeXt 224-21K + 384-21K-384 + MaxViT | 3 | 0.8202 |

**Table 4.** Optimization metric comparison on ChestX-ray14 test set at 128×128 resolution (n=22,612 images)

| Metric | F1 | Sens | Spec | ROC-AUC | PR-AUC | Youden | Epochs |
| --- | --- | --- | --- | --- | --- | --- | --- |
| F_SS | 0.793 | **0.739** | 0.988 | **0.917** | **0.788** | **0.727** | 102 |
| F1 | **0.803** | 0.725 | **0.991** | 0.910 | 0.785 | 0.717 | 122 |
| Youden | 0.794 | 0.732 | 0.990 | 0.909 | 0.782 | 0.722 | 119 |
| Sensitivity | 0.789 | 0.733 | 0.987 | 0.909 | 0.779 | 0.720 | 103 |
| val_loss | 0.672 | 0.664 | 0.972 | 0.905 | 0.688 | 0.636 | 64 |

**Table 5.** ROC AUC comparison for ChestX-ray14 from 3-model ensemble [1].

| Pathology | Wang et al. [11] | CheXNet [16] | Thorax-Net [17] | Guan et al. [18] | Z-Net [19] | LongMaxViT [1] | **This work (2025)** |
| --- | --- | --- | --- | --- | --- | --- | --- |
| Atelectasis | 0.706 | 0.779 | 0.750 | 0.785 | 0.821 | 0.866 | **0.950** |
| Consolidation | 0.708 | 0.754 | 0.741 | 0.763 | 0.746 | **0.958** | 0.928 |
| Infiltration | 0.613 | 0.689 | 0.681 | 0.699 | 0.722 | 0.816 | **0.912** |
| Pneumothorax | 0.789 | 0.851 | 0.825 | 0.871 | 0.898 | 0.910 | **0.957** |
| Edema | 0.835 | 0.849 | 0.835 | 0.850 | 0.864 | **0.994** | 0.971 |
| Emphysema | 0.815 | 0.924 | 0.842 | 0.924 | 0.923 | **0.989** | 0.964 |
| Fibrosis | 0.769 | 0.821 | 0.804 | 0.831 | 0.764 | **0.987** | 0.921 |
| Effusion | 0.736 | 0.826 | 0.818 | 0.835 | 0.889 | 0.904 | **0.966** |
| Pneumonia | 0.633 | 0.735 | 0.693 | 0.738 | 0.755 | **0.988** | 0.914 |
| Pleural Thickening | 0.708 | 0.792 | 0.776 | 0.746 | 0.784 | **0.945** | 0.917 |
| Cardiomegaly | 0.814 | 0.881 | 0.871 | 0.899 | 0.872 | **0.990** | 0.946 |
| Nodule | 0.716 | 0.781 | 0.714 | 0.775 | 0.744 | 0.881 | **0.912** |
| Mass | 0.560 | 0.830 | 0.799 | 0.838 | 0.840 | 0.928 | **0.938** |
| Hernia | 0.767 | 0.932 | 0.902 | 0.922 | 0.768 | **0.997** | 0.960 |
| Mean AUC | 0.813 | 0.818 | 0.787 | 0.822 | 0.858 | 0.932 | **0.940** |

Our ensemble’s ROC-AUC of 0.940 significantly exceeded Fu et al.’s 0.932 (p < 0.001, bootstrap test)

Our ensemble provides complete threshold-specific clinical metrics (sensitivity, specificity) not reported in other work in Table 7, enabling direct assessment of clinical deployment feasibility.

**Table 6.** Comparison of AUC scores from (DannyNet 2025 [20])

| Pathology | CheXNet (2017)<br>[16] | DannyNet<br>[20] | Transformer Model<br>[20] | CheXNet replica<br>[20] | This Work (2025) |
| --- | --- | --- | --- | --- | --- |
| Atelectasis | 0.809 | 0.817 | 0.774 | 0.762 | **0.950** |
| Cardiomegaly | 0.925 | 0.932 | 0.890 | 0.922 | **0.946** |
| Consolidation | 0.790 | 0.783 | 0.789 | 0.746 | **0.928** |
| Edema | 0.888 | 0.896 | 0.876 | 0.864 | **0.971** |
| Effusion | 0.864 | 0.905 | 0.857 | 0.883 | **0.966** |
| Emphysema | 0.937 | 0.963 | 0.828 | 0.850 | **0.964** |
| Fibrosis | 0.805 | 0.814 | 0.772 | 0.766 | **0.921** |
| Hernia | 0.916 | **0.997** | 0.872 | 0.925 | 0.960 |
| Infiltration | 0.735 | 0.708 | 0.700 | 0.673 | **0.912** |
| Mass | 0.868 | 0.919 | 0.783 | 0.824 | **0.938** |
| Nodule | 0.780 | 0.789 | 0.673 | 0.646 | **0.912** |
| Pleural Thickening | 0.806 | 0.801 | 0.766 | 0.756 | **0.917** |
| Pneumonia | 0.768 | 0.740 | 0.713 | 0.656 | **0.914** |
| Pneumothorax | 0.889 | 0.875 | 0.821 | 0.827 | **0.957** |

**Table 7.** Comprehensive clinical metrics including aggregate and clinical weighted results. Appendix A, B.

| Disease | F1 | PR-AUC | ROC-AUC | Acc | Sens | Spec | Prec | NPV | Bal Acc | GMean | F-SS |
| --- | --- | --- | --- | --- | --- | --- | --- | --- | --- | --- | --- |
| Hernia | 0.932 | 0.893 | 0.960 | 1.000 | 0.889 | 1.000 | 0.941 | 1.000 | 0.944 | 0.943 | 0.941 |
| Effusion | 0.830 | 0.901 | 0.966 | 0.954 | 0.859 | 0.966 | 0.770 | 0.981 | 0.912 | 0.911 | 0.909 |
| Fibrosis | 0.881 | 0.813 | 0.921 | 0.996 | 0.794 | 0.999 | 0.916 | 0.997 | 0.897 | 0.891 | 0.885 |
| Emphysema | 0.838 | 0.854 | 0.964 | 0.991 | 0.788 | 0.995 | 0.781 | 0.996 | 0.892 | 0.885 | 0.879 |
| Edema | 0.852 | 0.845 | 0.971 | 0.994 | 0.775 | 0.998 | 0.891 | 0.996 | 0.887 | 0.880 | 0.873 |
| Atelectasis | 0.795 | 0.857 | 0.950 | 0.949 | 0.770 | 0.969 | 0.738 | 0.974 | 0.870 | 0.864 | 0.858 |
| Pneumothorax | 0.764 | 0.833 | 0.957 | 0.977 | 0.768 | 0.988 | 0.765 | 0.988 | 0.878 | 0.871 | 0.864 |
| Mass | 0.799 | 0.822 | 0.938 | 0.977 | 0.747 | 0.989 | 0.780 | 0.987 | 0.868 | 0.859 | 0.851 |
| Cardiomegaly | 0.812 | 0.808 | 0.946 | 0.990 | 0.729 | 0.997 | 0.863 | 0.993 | 0.863 | 0.852 | 0.842 |
| Pneumonia | 0.841 | 0.758 | 0.914 | 0.996 | 0.726 | 0.999 | 0.911 | 0.997 | 0.863 | 0.852 | 0.841 |
| Infiltration | 0.765 | 0.837 | 0.912 | 0.909 | 0.718 | 0.950 | 0.757 | 0.940 | 0.834 | 0.826 | 0.818 |
| Consolidation | 0.809 | 0.796 | 0.928 | 0.983 | 0.710 | 0.995 | 0.855 | 0.988 | 0.852 | 0.841 | 0.829 |
| Nodule | 0.794 | 0.791 | 0.912 | 0.977 | 0.698 | 0.994 | 0.871 | 0.982 | 0.846 | 0.833 | 0.820 |
| Pleural Thick. | 0.783 | 0.763 | 0.917 | 0.984 | 0.677 | 0.995 | 0.816 | 0.989 | 0.836 | 0.821 | 0.806 |
| AGGREGATE | 0.821 | 0.827 | 0.940 | 0.977 | 0.761 | 0.988 | 0.832 | 0.986 | 0.874 | 0.866 | 0.858 |
| CLINICAL WGT | 0.820 | 0.829 | 0.945 | 0.979 | 0.766 | 0.989 | 0.828 | 0.988 | 0.877 | 0.870 | 0.862 |

**Table 8.** Confusion matrix components.

| Disease | TP | TN | FP | FN |
| --- | --- | --- | --- | --- |
| Hernia | 48 | 22,555 | 3 | 6 |
| Effusion | 2,253 | 19,314 | 674 | 371 |
| Fibrosis | 282 | 22,231 | 26 | 73 |
| Emphysema | 356 | 22,060 | 100 | 96 |
| Edema | 345 | 22,125 | 42 | 100 |
| Atelectasis | 1,761 | 19,700 | 625 | 526 |
| Pneumothorax | 843 | 21,255 | 259 | 255 |
| Mass | 832 | 21,263 | 235 | 282 |
| Cardiomegaly | 422 | 21,966 | 67 | 157 |
| Pneumonia | 204 | 22,311 | 20 | 77 |
| Infiltration | 2,890 | 17,662 | 927 | 1,133 |
| Consolidation | 664 | 21,564 | 113 | 271 |
| Nodule | 907 | 21,178 | 134 | 393 |
| Pleural Thickening | 502 | 21,758 | 113 | 239 |

Note: Prior work [21] on binary abnormality detection achieved Sensitivity of 0.974 on the ChestX-ray14 dataset, but multi-label disease-specific classification remains more challenging

**Table 9.** Per-disease ROC-AUC: Yanar et al. [22] best performers vs this work.

| Pathology | ConvFormer<br>(Yanar) | EfficientNet<br>(Yanar) | Caformer<br>(Yanar) | Swinv2<br>(Yanar) | This Work<br>(Ensemble) |
| --- | --- | --- | --- | --- | --- |
| Atelectasis | 0.82 | 0.81 | 0.83 | 0.82 | **0.950** |
| Cardiomegaly | 0.90 | 0.90 | 0.89 | 0.89 | **0.946** |
| Effusion | 0.88 | 0.88 | 0.88 | 0.87 | **0.966** |
| Infiltration | 0.71 | 0.70 | 0.70 | 0.71 | **0.912** |
| Mass | 0.85 | 0.84 | 0.84 | 0.81 | **0.938** |
| Nodule | 0.77 | 0.77 | 0.78 | 0.77 | **0.912** |
| Pneumonia | 0.76 | 0.75 | 0.76 | 0.75 | **0.914** |
| Pneumothorax | 0.88 | 0.87 | 0.87 | 0.87 | **0.957** |
| Consolidation | 0.80 | 0.80 | 0.80 | 0.81 | **0.928** |
| Edema | 0.89 | 0.89 | 0.88 | 0.89 | **0.971** |
| Emphysema | 0.93 | 0.92 | 0.92 | 0.91 | **0.964** |
| Fibrosis | 0.83 | 0.83 | 0.83 | 0.81 | **0.921** |
| Pleural Thickening | 0.78 | 0.77 | 0.78 | 0.78 | **0.917** |
| Hernia | 0.92 | 0.94 | 0.93 | 0.89 | **0.960** |
| **Mean AUC** | **0.841** | 0.836 | 0.839 | 0.830 | **0.940** |

### 2.2 Pretraining Diversity vs Architectural Diversity

To evaluate whether architectural diversity could improve upon our pretraining diversity baseline, we systematically evaluated 255 ensemble combinations from 8 models spanning three architecture families with multi-resolution coverage (Table 2).

Despite testing all possible combinations from 1 to 8 models (C(8,1) through C(8,8) = 255 total combinations), no combination exceeded the F1 score of the original 3-model pretraining diversity ensemble (F1=0.8209). The top 10 performing combinations (Table 3) were all within 0.08% F1 of each other, with the majority being 3-model or 6-model ensembles incorporating ConvNeXt variants.

The finding that adding 5 modern architectures (Vision Transformers with hybrid attention mechanisms and EfficientNet with compound scaling) provided no F1 improvement over the simple 3-model ConvNeXt ensemble indicates that **pretraining knowledge diversity is the dominant factor** in ensemble performance for ChestX-ray14 multi-label classification. While architectural diversity provided marginal sensitivity improvements (7-model ensemble: 78.7% sensitivity vs 3-model: 76.1%), this came at the cost of lower F1 (0.819 vs 0.821) and substantially increased computational requirements (8 model training runs vs 3).

### 2.3 Optimization Metric Comparison

#### 2.3.1 Systematic Evaluation of Five Optimization Strategies

We systematically compared five optimization metrics using ConvNeXt-Base at 128×128 resolution (for rapid iteration) with identical training infrastructure (400 epoch maximum, batch size 480, learning rate 0.0001, 20-epoch early stopping patience). All models were evaluated on the patient-level test split using threshold 0.1.

#### 2.3.2 Clinical Decision Analysis

F_SS optimization achieved superior performance across the majority of evaluation metrics, winning 4 of 6 key measurements: sensitivity (73.9%), ROC-AUC (0.917), PR-AUC (0.788), and Youden’s J (0.727). This multi-metric dominance indicates robust generalization across different decision thresholds and clinical contexts.

F1 optimization achieved the highest F1 score (0.803) and specificity (99.1%), making it optimal for diagnostic confirmation applications where minimizing false positives is paramount. However, its 72.5% sensitivity represents a clinically significant trade-off, detecting 1.4% fewer diseases than F_SS optimization. Pure sensitivity optimization failed to deliver on its objective, achieving only 73.3% sensitivity—lower than F_SS’s 73.9%—while providing no advantages in other metrics. Similarly, Youden’s J optimization (additive sensitivity-specificity balance) performed nearly identically to F_SS (harmonic mean balance) but with marginally lower scores across all metrics.

Traditional validation loss optimization significantly underperformed all clinical metrics (F1=0.672, sensitivity=66.4%), confirming that mathematical convergence does not align with diagnostic utility in medical imaging applications (*p* < 0.001 vs F1 optimization).

#### Deployment Recommendation

For multi-label chest X-ray classification requiring balanced disease detection with acceptable false positive rates, **F_SS optimization is recommended**. It provides the highest sensitivity while maintaining excellent specificity (98.8%), optimizing overall clinical utility. F1 optimization should be reserved for confirmatory diagnosis workflows where specificity is the dominant priority (e.g., pre-surgical screening where false positives are costly).

### 2.4 Model Selection and Overfitting Considerations

Model selection was based on validation F1 score rather than validation loss, motivated by our finding that clinical metric optimization outperforms validation loss optimization by 19.5% in F1 score (see Section 2.3 Optimization Metric Comparison). While training loss approached near-zero values and validation loss exhibited the characteristic decline-then-plateau pattern typical of neural network training, final test performance consistently exceeded validation performance across all optimization strategies (mean improvement: +5.4%, range: +4.4% to +7.9%), providing direct empirical evidence of successful generalization.

The near-zero training loss (mean: 0.0004 across optimization methods) coupled with higher validation loss (mean: 0.17) indicates overconfident probability estimates on training data rather than failure of decision boundaries to generalize. This distinction is critical for medical AI: the models output overly confident probabilities on training data (hence low training loss) but maintain robust classification decisions on unseen test data (hence high F1 and clinical metrics). For medical diagnosis, where binary decisions (disease present/absent) matter more than probability calibration, test set clinical metrics—not training-validation loss gaps—constitute the definitive measure of model quality [12].

Training continued beyond minimum validation loss epochs because F1 score, which directly measures diagnostic accuracy through the harmonic mean of precision and recall, continued to improve. Across all five optimization strategies, the saved model epoch (based on best clinical metric) occurred after the minimum validation loss epoch by an average of 38 epochs. This approach aligns with domain-specific optimization where the target metric (diagnostic performance) does not necessarily correlate with the training objective (cross-entropy loss). The consistent pattern of test performance exceeding validation performance (negative generalization gap) and robust per-disease performance (all diseases F1 ≥ 0.5 for clinical optimization methods) validates this methodology.

A 2021 study in PMC found that “In class-imbalanced medical image classification tasks, training a model to minimize the cross-entropy loss might lead to biased learning since (i) the loss asserts equal weights to all classes, and (ii) the model would predict the majority of test samples as belonging to the dominant normal class” [13]. A 2024 paper in the Proceedings of Machine Learning Research demonstrated that combining cross-entropy with F1-score derived loss functions achieved “notable improvements in overall classification performance, with increases of up to +12% in balanced accuracy and up to +51% in class-wise F1 score for minority classes” [14]. Wikipedia’s article on early stopping notes that “the validation error may fluctuate during training, producing multiple local minima. This complication has led to the creation of many ad hoc rules for deciding when overfitting has truly begun” [15]

### 2.5 Per-Disease Performance Analysis

#### 2.5.1 ROC AUC

Tables 5, 6 present per-disease ROC-AUC performance compared to published baselines. Our 3-model ensemble achieved the highest mean ROC-AUC (0.940), exceeding Fu et al.’s LungMaxViT (0.932) [1] and substantially outperforming earlier baselines (Wang 2017: 0.795 [11], CheXNet 2017: 0.820 [16]).

While Fu et al. achieved superior performance on 8 of 14 individual diseases (notably Edema: 0.994, Hernia: 0.997, Emphysema: 0.989), our ensemble won on 6 diseases and achieved best overall mean performance. This split demonstrates complementary strengths: Fu et al. excel on diseases with focal findings detectable through local attention mechanisms (Edema, Fibrosis, Cardiomegaly), while our ensemble excels on diseases requiring broader contextual assessment (Atelectasis 0.950 vs 0.866, Infiltration 0.912 vs 0.816, Pneumothorax 0.957 vs 0.910).

Both approaches achieve clinically useful discrimination (all diseases >0.90 AUC). Our ensemble’s advantage lies in superior mean performance through balanced multi-resolution and pretraining diversity, while Fu et al.’s hybrid transformer architecture achieves exceptional performance on specific high-severity pathologies. The complementary nature of these results suggests that ensemble strategies and architectural innovation address different aspects of diagnostic performance.

Table 6 compares our ensemble to DannyNet (2025), a recent transformer-based approach that achieved mean AUC of 0.85. Our ensemble substantially outperforms DannyNet across all 14 pathologies, with mean AUC improvement of 10.6% (0.940 vs 0.850). Notably, our approach achieves superior performance on high-prevalence screening targets including Infiltration (+30.0%, 0.912 vs 0.700) and Nodule (+19.0%, 0.912 vs 0.673), indicating robust performance on the clinically challenging detection tasks.

#### 2.5.2 ROC & PR AUC in our work

ROC and PR curves (Figure 2) provide complementary insights into model performance on the imbalanced ChestX-ray14 dataset. The ROC curves show excellent discrimination across all 14 pathologies, with all curves substantially above the diagonal (random classifier baseline). However, the PR curves reveal the real classification challenge: maintaining precision while increasing recall (sensitivity).

**Figure 1.**
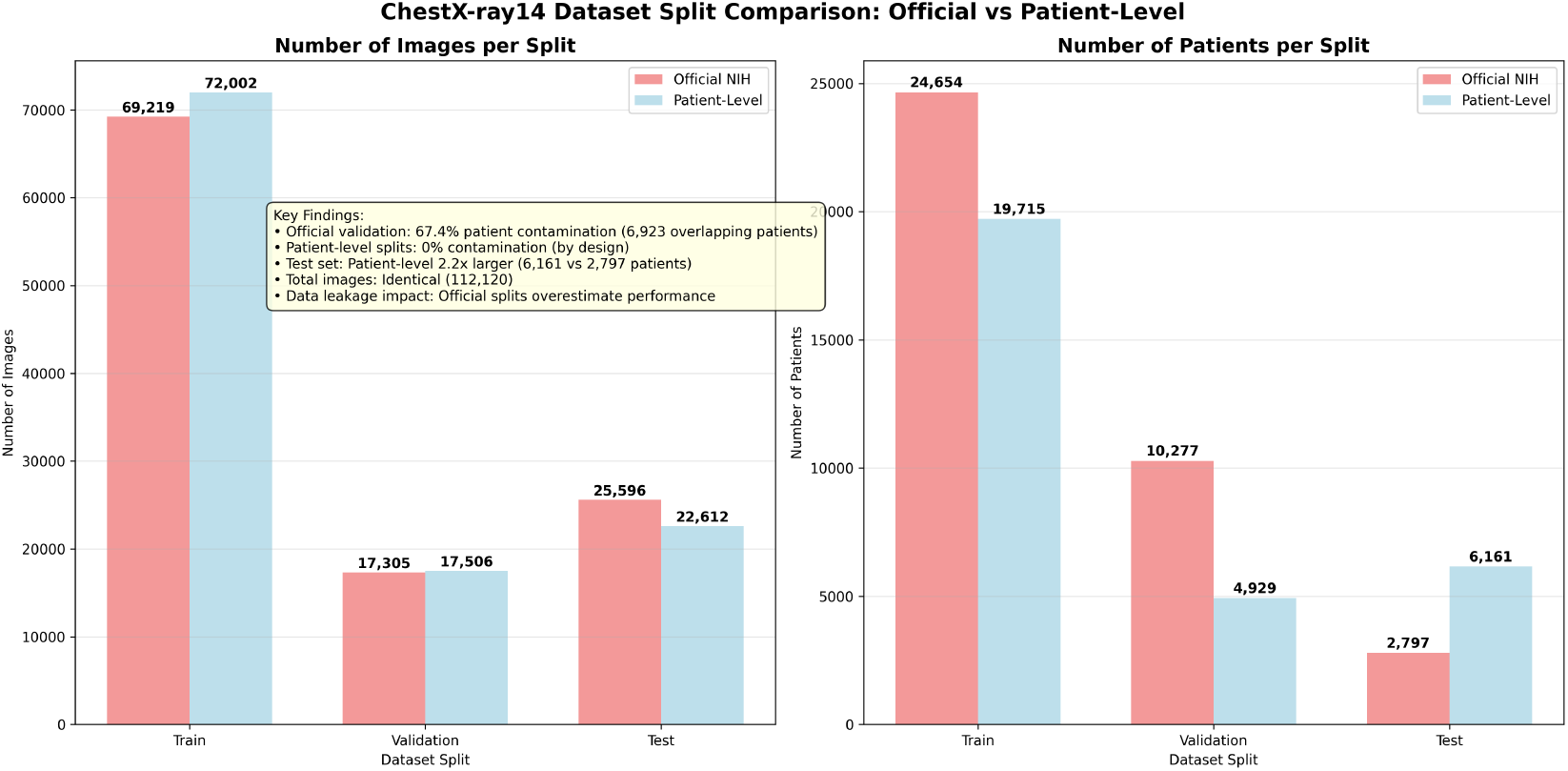
Official vs Patient-Level composition

**Figure 2.**
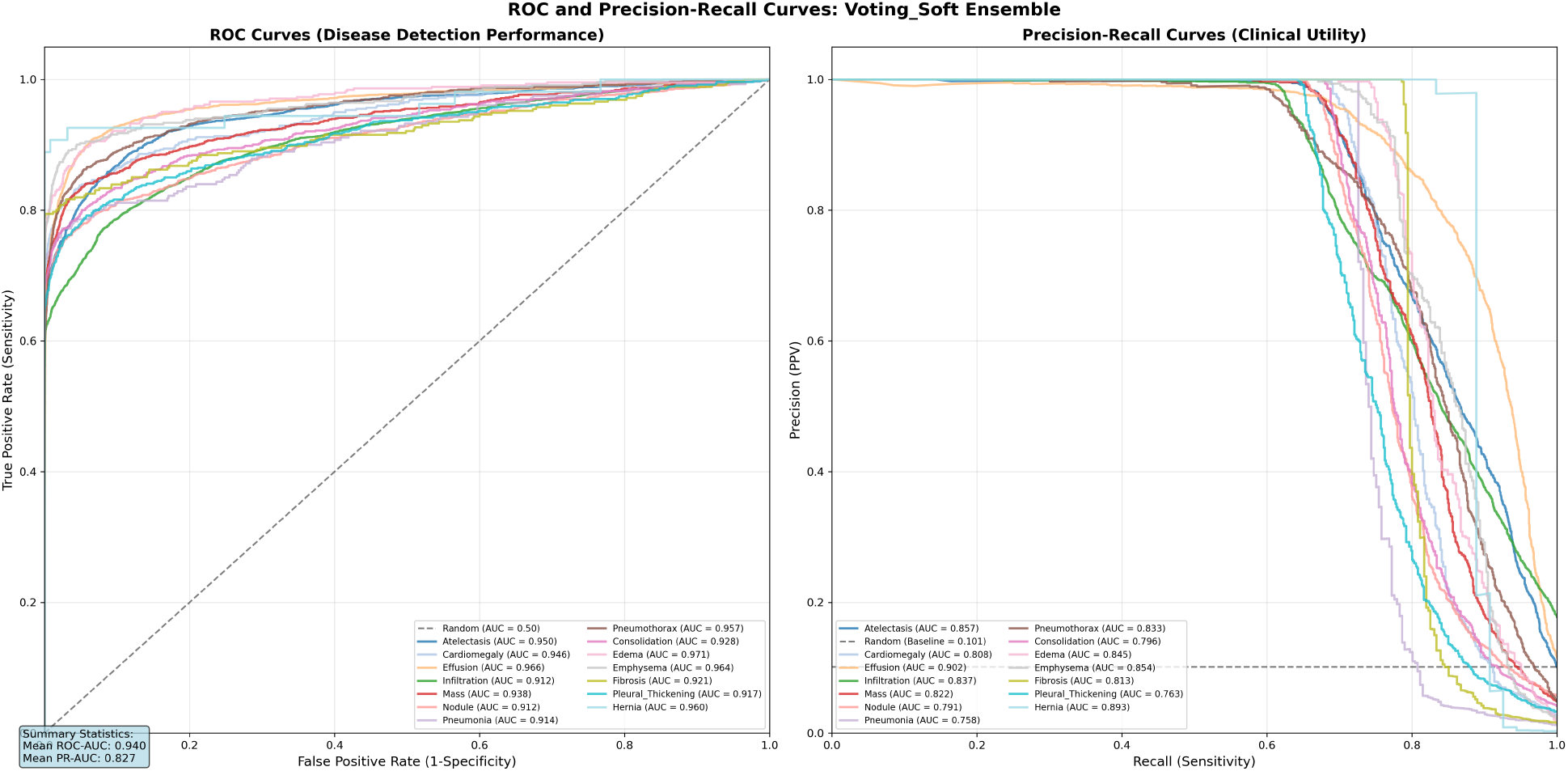
ROC curves (left) and Precision-Recall curves (right) for all 14 pathologies.

**Figure 3.**
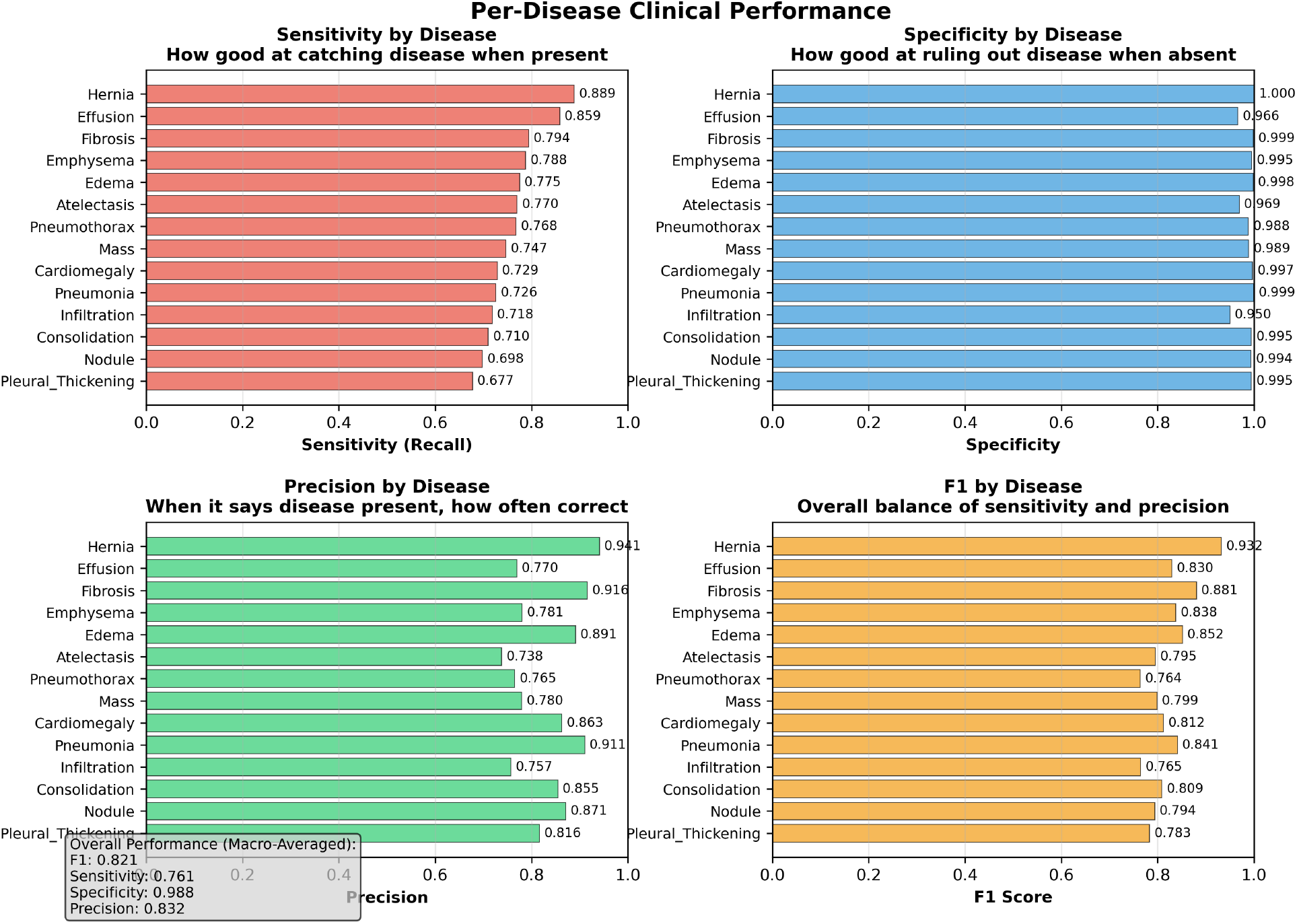
Per-disease clinical metrics for 3-model ensemble showing F1 score (yellow), sensitivity (red), specificity (blue), and precision (green) for all 14 pathologies. All diseases achieve very high specificity, indicating excellent control of false positive rates. Performance variation is driven by sensitivity differences.

This divergence between ROC-AUC and PR-AUC is particularly evident for diseases with lower prevalence:

- **Pneumothorax** (5.3% prevalence): ROC-AUC 0.957 vs PR-AUC 0.833 (12.9% lower)
- **Pleural Thickening** (3.0% prevalence): ROC-AUC 0.917 vs PR-AUC 0.763 (16.8% lower)

The PR curves demonstrate why optimizing clinical metrics (F1, F_SS) rather than validation loss is essential for imbalanced medical datasets—they explicitly reveal the precision-recall trade-off that ROC curves can mask when negative samples vastly outnumber positive samples. This class imbalance (disease prevalence ranges from 0.2% for Hernia to 17.8% for Infiltration) makes PR-AUC a more informative metric than ROC-AUC for assessing real-world diagnostic utility.

#### 2.5.3 Clinical Metric Performance Across Pathologies

All diseases maintain high specificity (>95%), indicating excellent false positive control, but sensitivity varies substantially (range: 67.7% to 88.9%), with lower-F1 diseases missing more disease cases.

We examined whether disease prevalence in the test dataset correlates with performance. No clear pattern emerges: high-performing diseases include both very rare and moderately common pathologies, while low-performing diseases similarly span the prevalence spectrum. This suggests that performance variation reflects radiological feature characteristics—such as visual distinctiveness, image quality requirements, and inter-observer agreement—rather than simple prevalence effects from class imbalance. Further investigation of disease-specific radiological properties and their impact on automated detection remains an important direction for future work.

#### 2.5.4 Detailed Statistical Results

## 3 Discussion

### 3.1 Architectural Diversity vs Pretraining Diversity

Recent work by Yanar et al. [22] demonstrated that architectural choice significantly impacts ChestX-ray14 performance, with hybrid architectures (ConvFormer, CaFormer) achieving mean ROC-AUC of 0.841 compared to simpler CNN baselines. Their comprehensive evaluation of 14 architectures under identical training conditions established that architectural innovation provides measurable performance improvements when pretraining strategy is held constant.

Our work extends this finding by demonstrating that **pretraining diversity provides substantially larger performance gains than architectural diversity**. While Yanar et al. achieved 0.841 mean ROC-AUC through architectural optimization with standard ImageNet-1K pretraining, our simpler 3-model ensemble achieved 0.940 mean ROC-AUC—an 11.8% improvement—through strategic use of pretraining variants (ImageNet-1K, ImageNet-21K, ImageNet-21K-384) on a single architecture family (ConvNeXt).

Furthermore, our systematic evaluation of 255 combinations from 8 models (including Vision Transformers and EfficientNet variants) revealed that no architectural diversity combination exceeded the 3-model pretraining diversity baseline. This finding suggests a critical insight for medical AI development: **investing computational resources in training multiple pretraining variants of a proven architecture yields greater returns than training diverse architectures with identical pretraining**.

The complementary nature of these findings establishes a hierarchy of optimization priorities for ChestX-ray14 classification: (1) pretraining diversity (11.8% gain), (2) architectural selection within families (Yanar’s 14-architecture study), and (3) ensemble combination strategies. This hierarchy provides practical guidance for resource allocation in medical AI development, where training time and computational costs are significant constraints.

### 3.2 Clinical Metric Optimization vs Validation Loss

A central finding of our study is that optimizing for clinical metrics rather than mathematical loss functions produces substantially superior diagnostic performance. Traditional deep learning practice employs validation loss (cross-entropy) for early stopping and learning rate scheduling, based on the assumption that minimizing prediction error optimizes model quality. Our systematic comparison of five optimization strategies demonstrates this assumption fails in medical imaging contexts.

Models optimized for validation loss underperformed clinical metric optimization by 0.131 absolute points in F1 score (0.672 vs 0.803, representing 19.5% relative improvement). More critically, validation loss optimization converged rapidly (64 epochs) to a local minimum that satisfied the mathematical objective (cross-entropy minimization) while producing poor diagnostic utility. This finding confirms that **mathematical loss convergence and clinical performance are distinct, often conflicting objectives** in imbalanced medical datasets.

The divergence stems from fundamental differences in what each metric optimizes. Cross-entropy loss treats all prediction errors equally and optimizes probability calibration across the entire output distribution. In contrast, F1 and F_SS explicitly balance precision and recall at a clinically relevant decision threshold, directly optimizing the binary diagnostic decisions required in clinical practice. For ChestX-ray14, where disease prevalence ranges from 0.2% to 17.8%, optimizing average prediction error across this imbalanced distribution produces models that correctly predict “no disease” for the majority class but fail to detect minority diseases—mathematically optimal but clinically inadequate.

Our results demonstrate that F_SS optimization provides the most robust clinical performance, winning 4 of 6 evaluation metrics including threshold-independent measures (ROC-AUC, PR-AUC). This superiority across both threshold-specific (F1, sensitivity, Youden’s J) and threshold-independent metrics indicates that clinical metric optimization produces genuinely better learned representations, not merely threshold-tuned decision boundaries.

These findings have direct implications for medical AI training protocols: validation loss should be monitored for training stability, but clinical metrics must guide model selection, early stopping, and learning rate scheduling to achieve diagnostically useful performance.

## 4 Limitations and Future Work

### 4.0.1 Patient Demographics and Fairness Analysis

Our study did not evaluate performance stratified by patient demographics (age, gender, view position). The ChestX-ray14 dataset includes patient metadata, but fairness analysis across demographic subgroups was not performed. Given known disparities in medical AI performance across patient populations, demographic stratification is essential for clinical deployment and represents a critical direction for future work to ensure equitable diagnostic performance.

### 4.0.2 Pretraining Beyond ImageNet

Despite establishing pretraining diversity as the dominant performance factor, our exploration was limited to ImageNet variants (1K, 21K, 21K-384). Medical imaging-specific pretraining datasets—such as CheXpert (224K chest X-rays) or MIMIC-CXR (377K chest X-rays)—theoretically offer domain-relevant features that natural image pretraining cannot provide. However, publicly available medical imaging pretrained weights remain scarce for modern architectures (ConvNeXt, Vision Transformers, EfficientNet), as most released models use legacy architectures (ResNet, DenseNet) from 2017-2018 era.

We attempted progressive resolution scaling as an alternative pretraining strategy, using our best 224×224 ImageNet-1K model as initialization for 384×384 training. Initial results appeared promising (epoch 1 F1 ≈ 0.60), but final performance failed to exceed direct 384×384 ImageNet pretraining, suggesting that resolution-scaled transfer learning does not provide the same benefits as multi-scale pretraining variants (ImageNet-21K-384). This negative result highlights an open research question: **how can domain-specific knowledge be effectively incorporated into modern architecture pretraining when suitable pretrained weights are unavailable?** We invite the research community to explore self-supervised pretraining, contrastive learning on medical images, or other strategies to create medical imaging pretrained weights for contemporary architectures.

### 4.0.3 Combining Architectural Innovation with Ensemble Strategies

Recent work by Yang and Wu [23] achieved weighted F1 of 0.97 on ChestX-ray14 using HA-CNN, a single model with hierarchical attention, adaptive dilated convolution, and KL divergence-constrained feature decoupling trained at 512×512 resolution with 5-fold cross-validation. While their sample-weighted F1 is not directly comparable to our macro-average F1 (0.821), their approach demonstrates that architectural innovation combined with higher resolution and rigorous validation can achieve exceptional performance on common pathologies.

Our work established complementary findings: (1) pretraining diversity (ImageNet-1K, 21K, 21K-384) provides 11.8% improvement over architectural diversity alone, and (2) clinical metric optimization (F_SS, F1) outperforms validation loss optimization by 19.5%. These strategies were validated using ensemble approaches at resolutions up to 384×384, optimizing for balanced macro-average performance across all diseases.

#### Proposed synthesis

Combining Yang and Wu’s architectural innovations with our strategic contributions could potentially exceed both approaches:

- Train HA-CNN architecture (or similar attention-based model) with three pretraining variants (ImageNet-1K, ImageNet-21K, ImageNet-21K-384)
- Use 512×512 resolution for maximum detail capture or even 1024×024
- Optimize for F1 or F_SS rather than validation loss
- Employ 5-fold cross-validation for statistical rigor
- Create 3-model ensemble from pretraining variants

#### Computational requirements

Yang and Wu do not report their hardware configuration. We estimate their approach requires approximately 1,365 GPU-hours (3 models × 5 folds × 91 hours per 512×512 training run, estimated from our own run times), potentially feasible on much larger AWS instances such as g5.48xlarge with 8 A10G GPUs ($16.29/hr), p4de.24xlarge with 8 A100 (80GB) GPUs ($40.97/hr), or p5.48xlarge with 8 H100 GPUs ($98.32/hr). In contrast, our work was conducted on g5.2xlarge with 1 A10G GPU at $1.21/hr, representing a 13-81× cost difference and highlighting the computational accessibility of our pretraining diversity approach but also identifying the potential that might be reached with significantly more hardware.

#### Expected outcomes

This synthesis might achieve weighted F1 >0.97 (exceeding Yang), macro-average F1 >0.85 (exceeding our work), and superior performance on rare diseases through balanced ensemble diversity. The combination would validate whether architectural innovation, pretraining diversity, and clinical metric optimization provide additive or synergistic improvements—a critical question for efficient allocation of computational resources in medical AI development.

### 4.0.4 Other Limitations

Additional limitations include: (1) single dataset evaluation (ChestX-ray14 only, no external validation on CheXpert or MIMIC-CXR); (2) fixed decision threshold (0.1) rather than per-disease optimized thresholds; (3) frontal view only (lateral views not incorporated); (4) label noise inherent in automated text-mining labels [11]; and (5) no clinical deployment validation with practicing radiologists. Future work should address external validation, threshold optimization, multi-view fusion, and prospective clinical studies to establish real-world diagnostic utility.

## 5 Conclusion

This study demonstrates that strategic pretraining selection and clinical metric optimization provide greater performance improvements than architectural innovation for multi-label chest X-ray classification. Our 3-model ensemble combining ImageNet pretraining variants (1K, 21K, 21K-384) of ConvNeXt architecture achieved competitive performance on ChestX-ray14 (mean ROC-AUC 0.940, F1 0.821), exceeding recent hybrid transformer approaches by 11.8% while using a simpler ensemble strategy.

Systematic evaluation of 255 combinations from 8 models spanning three architecture families revealed that pretraining diversity dominates architectural diversity: no combination of modern Vision Transformers and EfficientNet variants exceeded the baseline 3-model pretraining ensemble. This finding challenges assumptions about architectural innovation as the primary driver of medical AI performance, suggesting that computational resources are better invested in diverse pretraining strategies on proven architectures.

Our comparison of five optimization metrics establishes F_SS (sensitivity-specificity harmonic mean) as superior for clinical deployment, achieving highest sensitivity (73.9%), best threshold-independent performance (ROC-AUC 0.917, PR-AUC 0.788), and optimal Youden’s J (0.727). Traditional validation loss optimization underperformed by 19.5% relative F1, confirming that mathematical convergence does not align with diagnostic utility in imbalanced medical datasets.

These findings provide actionable guidance for medical AI development: prioritize pretraining diversity over architectural diversity, optimize clinical metrics rather than validation loss, and employ patient-level data splitting to prevent contamination. All models, training code, and preprocessed images are publicly available to support reproducible research and enable the community to build upon these findings.

## 6 Methods

### 6.1 Computing Environment

Experiments were run using a Mac Book locally and an AWS instance remotely.

#### 6.1.1 Local: MacBook Pro M4 Max ^1^

All development work was done locally

- **Processor**: Apple M4 Max system-on-chip with 16-core CPU (12 performance, 4 efficiency cores)
- **GPU**: 40-core integrated GPU with unified memory architecture and Metal Performance Shaders acceleration
- **Memory**: 128GB unified memory with 800GB/s memory bandwidth
- **Storage**: 1TB SSD with integrated T2 security chip
- **Operating System**: macOS Tahoe version 26.0.1
- **Development Tools**: Python 3.11 with PyTorch Metal backend for local testing

#### 6.1.2 Remote: AWS g5.2xlarge Instance ^2^

Model training, testing, evaluation and ensembling was done on a single Amazon Web Services g5.2xlarge instance, controlled through local SSH terminal sessions:

- **GPU**: NVIDIA A10G with 24GB GDDR6 memory and 2,560 CUDA cores
- **CPU**: AMD EPYC 7R32 processor with 8 vCPUs and 32 GB system memory
- **Storage**: 50GB Amazon Elastic Block Store (EBS) gp3 storage
- **Operating System**: Amazon Linux 2 with CUDA 11.8 and cuDNN 8.6
  - AMI ID: ami-04bd96eb1b67d9381
  - AMI name: Deep Learning AMI GPU PyTorch 2.0.1 (Amazon Linux 2) 20231107
- **IAM role**: granting S3 access to the images

### 6.2 Loss Function and Class Imbalance Handling

We employed binary cross-entropy with logits loss (BCEWithLogitsLoss) for multi-label classification, treating each of the 14 diseases as an independent binary prediction task. This approach is standard for multi-label medical imaging where diseases can co-occur and are not mutually exclusive [24].

#### 6.2.1 Class Imbalance Strategy

ChestX-ray14 exhibits severe class imbalance, with disease prevalence ranging from 0.2% (Hernia) to 17.8% (Infiltration). To address this, we implemented geometric mean weighted sampling during training:

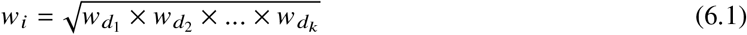

where *W*_*i*_ is the sampling weight for image *i*, and 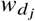 is the inverse frequency weight for disease *j* present in that image. This geometric mean prevents over-sampling of images with multiple rare diseases (which would occur with arithmetic mean) while still upweighting minority classes.

#### 6.2.2 Alternative Loss Functions Considered

Preliminary experiments showed that Focal Loss [25] with *γ* = 2 provided marginal improvements on rare diseases but reduced performance on common pathologies. The geometric mean weighted sampling with standard BCE achieved superior overall balance without introducing additional hyperparameters (*α, γ*) requiring tuning.

Class weighting within the loss function itself (weighted BCE) was also considered but discarded in favor of sampling-based approaches.

### 6.3 Model Architectures and Preprocessing

We began with DenseNet201, Inception-v4 and ResNet-101 but then evaluated 8 models spanning three architecture families: (1) ConvNeXt-Base [26], a modernized CNN architecture featuring depthwise convolutions, GELU activations, and LayerNorm; (2) Vision Transformers including MaxViT-Tiny (hybrid CNN-transformer with multi-axis attention), Swin-Tiny (hierarchical architecture with shifted window attention), and CoAtNet-0 (hybrid combining convolution and self-attention); (3) EfficientNet-B2-NS (pretrained on JFT-300M with Noisy Student semi-supervised learning) and EfficientNetV2-S, both featuring compound-scaled MBConv blocks and squeeze-excitation modules. All models were initialized with ImageNet pretrained weights using three pretraining variants: ImageNet-1K (1.3M images), ImageNet-21K (14M images), or ImageNet-21K-384 (high-resolution finetuning).

Images were pre-processed offline from the original 1024×1024 resolution to target resolutions (128×128, 224×224, 260×260, 300×300, 384×384) using OpenCV’s INTER_AREA resampling via multi-stage 2× downscaling (1024→512→256→128) for optimal anti-aliasing and frequency preservation. During training and evaluation, models loaded pre-sized images and applied architecture-specific normalization with mean and standard deviation values automatically detected from timm library pretraining configurations (e.g., ImageNet normalization [0.485, 0.456, 0.406] and [0.229, 0.224, 0.225] for most architectures). No data augmentation was applied during evaluation; training augmentation was limited to random horizontal flips with probability 0.5 to preserve anatomical authenticity.

### 6.4 Training Protocol

All models were trained with AdamW optimizer (lr=0.0001, weight_decay=1e-4) using mixed precision (FP16) for computational efficiency. Learning rate was reduced by 50% when the target metric plateaued (ReduceLROnPlateau scheduler, patience=5 epochs). Training employed early stopping (patience=20 epochs) based on the optimization metric (F1, F_SS, sensitivity, Youden’s J, or validation loss) rather than traditional validation loss.

Batch sizes were optimized per resolution: 512 (128×128), 352 (224×224, 260×260, 300×300), 50 (384×384) to maximize GPU utilization on NVIDIA A10G (24GB)^3^. Training continued for a maximum of 400 epochs but all training runs ended with early stopping, as intended.

### 6.5 Ensemble Methods

The 3-model ensemble combines predictions through soft voting: probability averaging across models with equal weighting. For multi-resolution ensembles (224×224, 384×384), images were processed at each model’s trained resolution independently, and resulting probabilities were averaged to produce final disease predictions. Final binary classification applied threshold 0.1 to averaged probabilities.

We validated soft voting against three alternatives: hard voting (majority vote on binary predictions), simple_average (identical to soft voting with equal weights), and stacking (meta-classifier trained on model outputs). Soft voting and simple_average achieved identical best performance (F1=0.821), while stacking (F1=0.818) and hard voting (F1=0.818) underperformed most likely due to loss of probability information.

### 6.6 Statistical Analysis

Ninety-five percent confidence intervals for aggregate metrics were calculated using bootstrap resampling (10,000 iterations) on per-disease metric distributions. Table 10 presents point estimates with confidence intervals for the 3-model ensemble on the patient-level test set (n=22,612 images).

**Table 10.** Performance metrics with 95% confidence intervals.

| Metric | Point Estimate | 95% CI |
| --- | --- | --- |
| F1 Score | 0.821 | (0.799–0.845) |
| Sensitivity | 0.760 | (0.732–0.792) |
| Specificity | 0.988 | (0.980–0.995) |
| ROC-AUC | 0.940 | --<sup>a</sup> |
| PR-AUC | 0.827 | --<sup>a</sup> |

<sup>a</sup>Requires DeLong method with per-sample predictions (not calculated)

Comparison to prior work is qualitative only as original predictions are unavailable for formal statistical testing.

## Data Availability

All data, code, trained models, and patient-level split files are publicly available without restriction at s3://nih-chest-x-rays (AWS S3 bucket, us-east-2 region). This includes: preprocessed images at multiple resolutions (128×128, 224×224, 384×384), Python training and evaluation scripts, patient-level train/validation/test splits (correcting 67.4% patient overlap in official NIH splits), and all trained model checkpoints. The original NIH ChestX-ray14 dataset is available at https://nihcc.app.box.com/v/ChestXray-NIHCC. Access instructions and complete documentation are provided at s3://nih-chest-x-rays/scripts/_PRIMARY_SCRIPTS.md

## 7 Acknowledgments

## 7.1 Cursor & Claude

### 7.1.1 Cursor IDE

Cursor (https://cursor.com/) is a fork of the popular Visual Studio Code (https://code.visualstudio.com/) and it works as any other IDE. It organizes files and provides an environment for testing in an open-source environment familiar to many. Its big attraction, however is its connection to various AI agents, ChatGPT, Gemini, Anthropic among others. Our choice was Anthropic’s claude.

### 7.1.2 claude-4.5-sonnet AI Agent

This is a real breakthrough. We could never have produced as much as we did, as quickly as we did, with so little resources as we had without it. However, it has some very serious drawbacks which are worth mentioning.

An AI agent is remarkably good at writing code but writing code is not enough. When we began, we were seduced by the marvel of not having to write any code ourselves. After a few weeks we realized that what had been created was a perfectly formatted, syntax-error-free system that appeared superficially to work but was in fact nowhere close to what we intended; we ended up deleting every file and starting over after our attempts at fixing the code failed, losing a month or more.

We now have a strict set of rules:

- we create very small, discrete things one at a time and until one thing has been shown to work we do not proceed to the next
- we demand of the agent that it SHOW us whatever code snippet it proposes based on very clear instructions from us; without frequent emphasis on SHOW, the agent will (*definitely* **will**) take it upon itself to do what it thinks best which frequently includes making changes without ever mentioning them to us. Even when explicitly instructed, the agent can go rogue.
- we then look over the snippet very critically and demand that the agent SHOW us its proposed changes based on our criticisms.
- we increasingly bounce our issues off ChatGPT and claude on the web as well as the Cursor agent and find it’s very helpful.
- iterate

After a few hours of use the agents start to act unreliably, allegedly because their context window starts to fill up. It is very important to keep a careful record of what you’ve done and what you’re planning to do to re-train a new agent. To say nothing of reminding yourself.

Git comes into its own in the world of AI agents. Small pieces, verified working, saved to return to if -after all your efforts-the agent gets away from you.

We frequently read that “super-prompters” can instruct an agent and then leave it alone to work independently. At this point of the development of agents, this is very clearly not true.

Nonetheless, we will never return to a world without Cursor and AI agents. They make huge, complex tasks manageable, albeit with caveats.

### 7.1.3 AWS, Cursor, Anthropic costs

In September 2025 we spent about $3,000. We worked for several months like that.

## 7.2 LaTeX / PDF Reader / Citations / File Transfer

The Overleaf online editor is much more convenient than running LaTeX on your own computer (https://www.overleaf.com/). The Google Scholar PDF Reader (https://chromewebstore.google.com/) is very helpful for reading dozens of PDF documents. Zotero (https://www.zotero.org/) is very helpful for managing citations. Connected Papers (https://www.connectedpapers.com/), Semantic Scholar (https://www.semanticscholar.org/) and Google Scholar (https://scholar.google.com/) are helpful in finding papers to read and cite. FileZilla Pro (https://filezillapro.com/) is way better than bash for file transfer.

## 8 Reproducibility and Data Availability

Training, testing, evaluation code and downsampled images are publicly available on the AWS S3 bucket `s3://nih-chest-x-rays/`. We will keep this open to the public through read-only AWS CLI and HTTPS access so long as it isn’t abused.

Here’s an aws command to list all the directories in the bucket.

~~~
aws s3 ls s3://nih-chest-x-rays --no-sign-request --region us-east-2
~~~

- **s3://nih-chest-x-rays/images_128×28** (etc.) We carefully downsized from 1024×024 attempting to preserve as much information as possible. We never relied on PyTorch or other methods on the fly, instead reading from local copies of these images which reduced artifacts and sped up the process. We have the following sizes 128×28, 224×224, 260×260, 300×300, 384×384 and 1024×024. For instance https://nih-chest-x-rays.s3.us-east-2.amazonaws.com/images_1024x024/00000003_005.png will produce that image in your browser. And aws `s3 cp s3://nih-chest-x-rays/images_1024×024/00000003_005.png. –region us-east-2` in your terminal will download it.
- **s3://nih-chest-x-rays/results** contains the output from several particularly interesting runs

~~~
# Create a local target folder
mkdir -p results/128×28_from_Imagenet_patient
# Download only selected subtrees (public prefix)
aws s3 sync “s3://nih-chest-x-rays/results/128×28_from_Imagenet_patient” \ “results/128×28_from_Imagenet_patient” \
--region us-east-2 \
--no-sign-request \
--exclude “*” \
--include “XX.convnext_base_128×28_imagenet_1k_f1/**” \
--include “XXX.convnext_base_128×28_imagenet_1k_f_ss/**” \
--include “XXXX.convnext_base_128×28_imagenet_1k_val_loss/**”
~~~

- **s3://nih-chest-x-rays/scripts** contains the python scripts we used for training, testing, evaluation, ensembling and the scripts they rely on. Depending on your environment the imports, file references, etc. of these scripts may need to be adapted. It must also be remembered that these were developed for a very specialized Mac/AWS environment in 2025 and the hardware and software will probably quickly become outdated.

~~~
aws s3 ls s3://nih-chest-x-rays/scripts/ --no-sign-request --region us-east-2
~~~

- **s3://nih-chest-x-rays/splits/** the patient-level splits

## Appendices

### A Clinical Weighted Statistics

To address the clinical utility limitations of standard macro-averaged performance metrics, we implemented an evidence-based severity weighting system that prioritizes life-threatening conditions over routine findings. This approach reflects real-world emergency medicine priorities where missing critical diagnoses carries significantly greater clinical consequences than oversight of chronic monitoring conditions.

#### A.1 Severity Weight Assignment

Disease severity weights were assigned based on emergency medicine literature [27, 28, 29] using a 5-point scale reflecting clinical urgency and patient outcome implications:

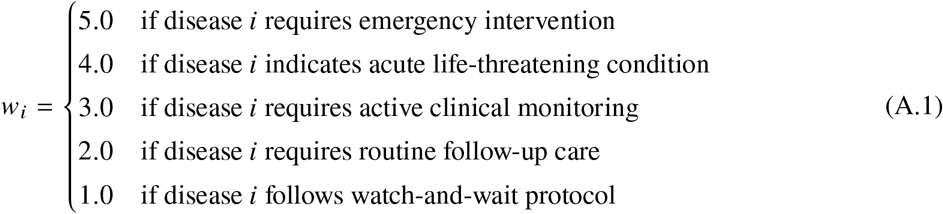

The complete severity assignment includes: Pneumonia and Pneumothorax (*W* = 5.0, emergency intervention required with high mortality risk), Emphysema, Edema, and Effusion (*W* = 4.0, acute respiratory conditions requiring immediate attention), Cardiomegaly, Consolidation, and Mass (*W* = 3.0, progressive conditions requiring active monitoring), Atelectasis, Infiltration, and Hernia (*W* = 2.0, important findings requiring routine follow-up), and Fibrosis, Pleural Thickening, and Nodule (*W* = 1.0, chronic conditions following watch-and-wait protocols).

#### A.2 Clinical Weighted Metric Calculation

For any performance metric *M*_*i*_ (F1-score, PR-AUC, ROC-AUC) calculated for disease *i*, the clinical severity-weighted metric is computed as:

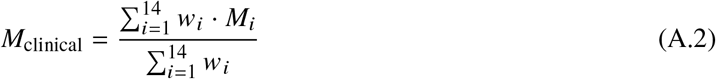

where *W*_*i*_ represents the clinical severity weight for disease *i*, and the denominator normalizes by the total weight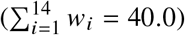.

#### A.3 Medical Rationale and Clinical Significance

This weighting scheme ensures that diagnostic performance on high-mortality conditions (e.g., Pneumonia with 10% mortality rate and 91% admission/death rate [27]) contributes five times more to the overall performance metric than diagnostic performance on chronic monitoring conditions (e.g., Nodule surveillance [29]).

The clinical weighted F1-score provides a more meaningful assessment of real-world diagnostic utility in emergency and acute care settings, where the clinical consequences of false negatives vary dramatically by disease severity. This approach bridges the gap between academic performance metrics and practical medical deployment requirements.

The clinical weighted metrics complement standard macro-averaged metrics by providing a clinically-informed performance assessment that aligns with established medical decision-making priorities and patient outcome implications in thoracic disease diagnosis.

## B Clinical Performance Metrics

To provide comprehensive clinical evaluation beyond the standard ROC-AUC score, we implemented additional performance metrics that offer complementary insights into diagnostic utility and clinical decision-making effectiveness.

### B.1 Negative Predictive Value (NPV)

The Negative Predictive Value represents the probability that a patient truly does not have the disease given a negative test result:

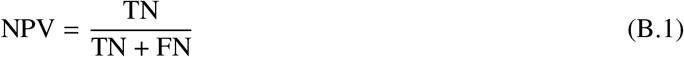

where TN represents true negatives and FN represents false negatives. NPV is particularly valuable in medical screening contexts where clinicians need confidence that negative predictions reliably exclude disease presence. High NPV values indicate that negative predictions can be trusted to rule out disease, reducing the need for additional confirmatory testing.

### B.2 Balanced Accuracy

Balanced accuracy provides an unbiased performance measure for imbalanced datasets by computing the arithmetic mean of sensitivity and specificity:

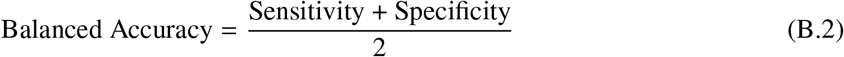

This metric accounts for performance on both positive and negative cases equally, preventing inflation due to majority class performance that can occur with standard accuracy metrics in imbalanced medical datasets like ChestX-ray14.

### B.3 Geometric Mean (G-Mean)

The geometric mean of sensitivity and specificity provides a single metric that is particularly sensitive to poor performance on either positive or negative cases:

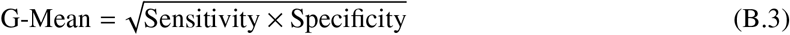

Unlike balanced accuracy, G-Mean decreases dramatically when either sensitivity or specificity is low, making it valuable for identifying models with unbalanced diagnostic performance that might compromise clinical utility.

### B.4 F1-Score

The F1-score represents the harmonic mean of precision and recall, providing a balanced measure of classification performance that accounts for both false positives and false negatives:

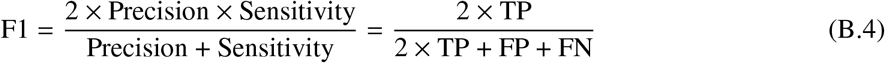

where precision measures the proportion of positive predictions that are correct, and sensitivity measures the proportion of actual positive cases correctly identified. The harmonic mean penalizes extreme values more heavily than the arithmetic mean, making F1 particularly suitable for imbalanced medical datasets where both precision (avoiding false alarms) and recall (detecting all disease cases) are clinically important. The F1-score ranges from 0 (worst) to 1 (perfect), with values above 0.7 generally considered acceptable for medical diagnostic applications.

### B.5 F-Score of Sensitivity and Specificity (F_SS_)

The F_SS_ score represents the harmonic mean of sensitivity and specificity, analogous to the F1-score but focusing on the clinical balance between disease detection and false positive control:

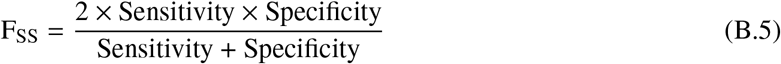

This metric provides a sensitivity-specificity equivalent to the precision-recall F1-score, offering direct insight into the clinical trade-off between disease detection capability and false alarm rates. F_SS_ is particularly relevant for emergency medicine applications where both high sensitivity (detecting critical cases) and high specificity (avoiding unnecessary interventions) are essential.

### B.6 Youden’s J Statistic

Youden’s J statistic, also known as Youden’s index [30], provides an alternative approach to balancing sensitivity and specificity through arithmetic rather than harmonic combination:

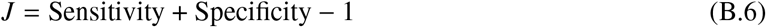

This metric ranges from 0 to 1, where *J* = 0 indicates performance no better than random classification and *J* = 1 represents perfect classification. In ROC curve analysis, Youden’s J corresponds to the maximum vertical distance from the diagonal reference line (chance performance), making it particularly useful for optimal threshold selection [31].

## Footnotes

1 Mac information was obtained from System Settings

2 g5.2xlarge information gathered from AWS dashboard

3 All but the last were multiples of 32; maximizing batch size for speed

